# Effects of Short and Intensive Bimanual Training on Spatiotemporal Characteristics of Bimanual Coordination and Motor Learning in Children with Unilateral Cerebral Palsy

**DOI:** 10.1101/2025.08.29.25334738

**Authors:** Shailesh S. Gardas, John Willson, Swati M. Surkar

**Affiliations:** Department of Physical Therapy, East Carolina University, Greenville, NC 27834, United States

**Author notes:** Corresponding author Swati M. Surkar Department of Physical Therapy East Carolina University, Greenville, NC 27834, USA.

**Keywords:** Unilateral cerebral palsy, kinematics, bimanual coordination, training intensity, motor function

## Abstract

**Objectives:** This study aimed to compare the effects of short versus intensive bimanual training on spatiotemporal features of bimanual coordination in children with unilateral cerebral palsy (UCP).

**Methods:** In a prospective, repeated-measures design, 28 children with UCP completed two training regimens: a short dose (75 repetitions of speed stacking; ∼1–1.5 hours) and an intensive dose (30 hours of Hand arm bimanual intensive training). Bimanual learning was indexed by average time to complete nine stacking trials. Spatiotemporal kinematics were evaluated using three-dimensional motion analysis. For the bimanual coordination task, (3–2–1 stacking) outcomes included normalized movement overlap, total task duration, and participation time. For a symmetric bimanual task (simultaneous two-cup transfer), task synchronization and completion time were analyzed. Peak tangential velocity and hand trajectory were assessed across both tasks. General linear models with repeated measures were used to analyze the effects of training dose and extremity.

**Results:** There was a significant main effect of training dose on movement time (p < 0.001), with both doses improving bimanual learning. The intensive dose yielded significantly greater gains in normalized movement overlap, total task duration, hand trajectory, and participation time (all p = 0.001) during the bimanual coordination task. A dose-by-extremity interaction was identified for peak tangential velocity (p = 0.03), demonstrating greater velocity gains in the more affected limb. In the symmetric task, a main effect of dose was found only for hand trajectory (p < 0.03).

**Conclusions:** Both short and intensive bimanual training enhanced bimanual learning and coordination in children with UCP. While intensive training yielded greater improvements, even brief, ecologically valid tasks produced measurable gains, highlighting the importance of training intensity and task specificity in pediatric neurorehabilitation.

## 1. Introduction

Children with unilateral cerebral palsy (UCP) exhibit sensorimotor impairments such as weakness, spasticity, dyskinetic movements, and motor planning deficits, resulting in abnormal motor control on one side of the body (Surkar et al., 2019; Surkar, Hoffman, Davies, et al., 2018; Surkar, Hoffman, Harbourne, et al., 2018). These impairments hinder the child’s ability to perform coordinated bimanual tasks such as tying shoelaces, buttoning, eating, etc. which are essential daily activities required for functional independence (Birtles et al., 2011; Coluccini et al., 2007; Y.-C. Hung et al., 2012). Moreover, due to sensorimotor impairments, children with UCP often rely on their less affected upper extremity (UE) and do not actively engage the preserved capacity of their more affected UE in bimanual tasks (Houwink et al., 2011). This limited use of the more affected UE can lead to developmental disregard, where the child becomes less aware of their affected hand, exacerbating bimanual coordination difficulties over time impacting functional independence and overall quality of life (Steenbergen et al., 2008; Zielinski et al., 2014).

Intensive motor-learning interventions, such as Hand-Arm Bimanual Intensive Therapy (HABIT) (Gardas et al., 2023; Surkar, Hoffman, Willett, et al., 2018) and Constraint-Induced Movement Therapy (CIMT) (Dong et al., 2013), have shown promise in improving bimanual coordination and enhancing the function of the more affected UE in children with UCP. Common clinical assessments such as the Jebsen Hand Function Test, Nine-Hole Peg, Assisting Hand Assessment (AHA), and Canadian Occupational Performance Measure (COPM) have been used to evaluate effectiveness of these interventions. (Ouyang et al., 2020; Shierk et al., 2016). While these assessments are important, they have notable limitations as they often focus on unimanual dexterity or the spontaneous use of the more affected UE in bimanual tasks, but they lack the ability to objectively measure the quality of hand use, movement accuracy, or the spatiotemporal characteristics of both hands during bimanual activities (Gardas et al., 2023; Jaspers et al., 2009). Furthermore, some of these assessments such as AHA require expert coding based on video recordings, while others like COPM, are prone to subjective biases (Adams, 2005; Elad et al., 2013). To better capture the dynamic and complex nature of bimanual coordination, more objective and comprehensive measures are needed (Jaspers et al., 2009).

Three-dimensional movement analysis (3DMA) offers an objective method for quantifying spatiotemporal characteristics of bimanual coordination during real-life tasks (Jaspers et al., 2009). While 3DMA has been used to study unimanual tasks or basic bimanual tasks such as reaching and grasping, it has rarely been applied to functional activities that more accurately reflect daily life (Kelso et al., 1979; Steenbergen et al., 1996; Swinnen, 2002). Recent studies have incorporated 3DMA for bimanual tasks like drinking from a glass, moving a box on a desk, reaching for a light switch or child-friendly protocols, such as the Nintendo Wii, or gaming scenarios requiring both hands to fly a plane with a joystick (Cacioppo et al., 2020; Klotz et al., 2014; Mailleux et al., 2017; Rudisch et al., 2016). However, many of these protocols are either too simple to fully capture coordination patterns or involve complex experimental setups that limit feasibility. Tasks like cup stacking, which require coordination of both hands to complete complex, multi-step movements, represent a more ecologically valid measure of bimanual coordination. (Surkar et al., 2023) Cup stacking requires reaching, grasping, moving, and releasing cups in a specific pattern, demanding coordination of both hands. Cup stacking involves both in-phase and anti-phase movements (Kelso & Schöner, 1988; Swinnen, 2002), engaging the spatiotemporal coordination required for many daily activities. This study employed two distinct cup stacking tasks to investigate the kinematics of both in-phase and anti-phase movements. The symmetric performance task, in which children rapidly pick up and place cups into a target using both hands, was designed to assess in-phase movement. In contrast, a coordination task, involving the stacking of six cups in a specific pattern through bilateral cooperation, was utilized to evaluate anti-phase movements.

Another important aspect of this study is the comparison of spatiotemporal characteristics of bimanual coordination following a short-bout and intensive-bout of bimanual training. Historically, studies assessing spatiotemporal characteristics of bimanual coordination in children with UCP have focused on intensive dose (90 hours) of bimanual therapy (Ouyang et al., 2020). Despite the promise of intensive interventions, these intensive therapies can be costly and time-consuming. The typical therapy schedule for children with UCP is 1–2 hours of therapy per week over several months (Ferrante et al., 2019), raising the question of how even brief periods of practice might impact bimanual coordination. Previous studies, such as those involving the drawer-opening task in the HABIT protocol, have demonstrated improvements in spatiotemporal measures of bimanual coordination with intensive therapy (Y. C. Hung et al., 2011). However, no research has specifically compared these improvements to shorter duration (1-1.5 hours) training bouts. This study aims to fill this gap by investigating the effects of short- and intensive bouts of bimanual training, designed to mirror the typical duration of clinical therapy session. By analyzing spatiotemporal characteristics with 3DMA after this brief bout of bimanual practice, we aim to gain valuable, objective insights into changes in movement accuracy, overlap, and the trajectory of the more affected UE that could be missed by traditional assessments, such as the AHA.

Therefore, this study aims to investigate how different doses (short- vs. intensive bout) of bimanual training affect the spatiotemporal characteristics of bimanual coordination in children with UCP. We hypothesize that both short and intensive bimanual training will improve coordination, as reflected in spatiotemporal metrics of the coordination cup stacking task, with greater gains following the intensive as compared to the short bout of training. By addressing existing gaps in both clinical assessment methods and intervention practices, this research will contribute valuable information on how different bouts of bimanual practice can influence functional outcomes in daily tasks, potentially offering a more feasible and accessible approach for improving bimanual coordination in this population.

## 2. Methods

### 2.1 Trial Design

This study is an ancillary analysis of randomized controlled trial (NCT05355883 and NCT05777070). It was a prospective repeated measures training study conducted at the Pediatric Assessment and Rehabilitation Laboratory (PeARL) and Human Movement Analysis Laboratory (HMAL) at East Carolina University (ECU), NC. The University and Medical Center Institutional Review Board, ECU, approved the study. We obtained written parental consent and child assent. The study was conducted between November 2021 and January 2024.

### 2.2 Participants

This study included 28 participants with UCP, who were within 6 – 16 years of age, Manual Ability Classification System (MACS) levels I-III, able to complete a stack of three cups within one minute, and sufficient cognition to follow the experiment instructions. Children with other neuromotor disabilities, cognitive and communication deficits, cardiorespiratory dysfunctions, metabolic disorders, neoplasms, and a history of botulinum neurotoxin injections on the affected UE, and history of intensive UE therapy (HABIT or CIMT) in the past 6 months were excluded.

### 2.3 Procedures

**Figure 1.**
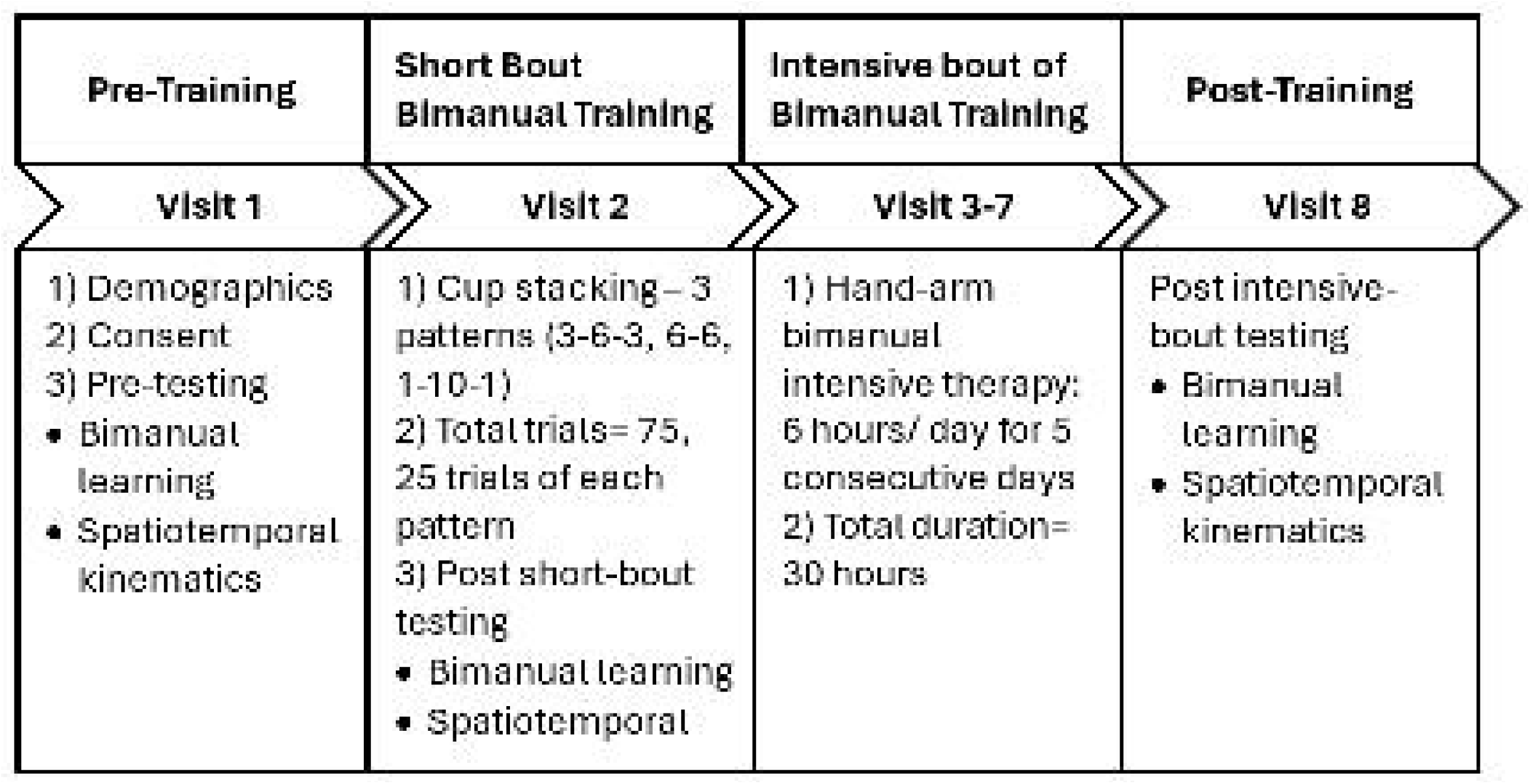
Schematics of study protocol.

### 2.4 Interventions

#### 2.4.1 Short bout of Bimanual Training Using Speed Stack

The short bout of bimanual training involves a cup stacking task developed by the World Sport Stacking Association. This task requires grasping, moving, and releasing cups, necessitating both hands to work in coordination, simulating upper extremity skills that are often trained during rehabilitation. Additionally, the cup stacking task was selected because it has been used successfully to assess motor learning and bimanual coordination in children with UCP, healthy adults, and post-stroke survivors (Surkar et al., 2023; Tretriluxana et al., 2015; Udermann et al., 2004).

The short bout training session consisted of three cup stacking patterns (3–6–3, 6–6, and 1–10–1; see Figure 2) using 12 specialized cups designed for speed stacking (Speed Stacks, Castle Rock, CO, USA) (Surkar et al., 2023). Participants practiced for 75 trials of cup stacking, divided into three blocks of 25 trials for each pattern. On average, the whole training session lasted for 1 – 1.5 hours including rest pauses. Two to five minutes of rest breaks were provided after every 15 trials. In each block, the sequence of practice patterns was randomized, and the experimenter informed the participants about their performance or movement time after each trial.

**Figure 2.**
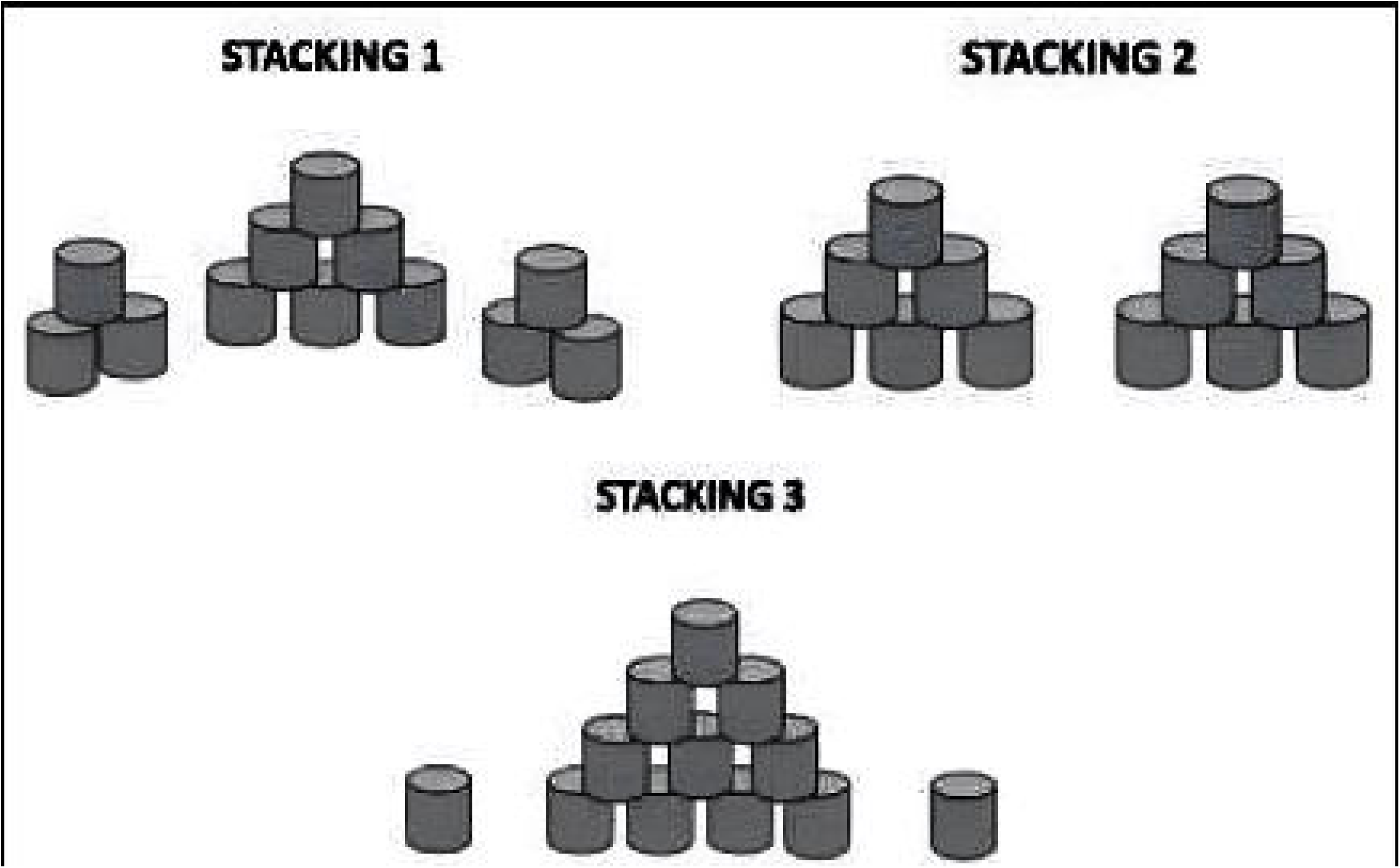
Stacking patterns for short bout of bimanual practice. Children practiced a total of 75 trials of cup stacking during visit 2 of the study (25 trials each for three patterns).

Before starting each trial, the cups were placed upside down and nested on the mat according to the pattern being practiced. For example, when practicing the first pattern, the cups were arranged with three on the left, six in the center, and three on the right. The experimenter demonstrated the task, and participants performed three practice trials to become familiar with the task and the patterns. Specific instructions were provided for the participants: 1. go as fast as possible; 2. always use both hands; 3. stack the cups first and then unstack; 4. stacking and unstacking must be done from left to right; 5. if errors occur, fix them and continue stacking; and 6. the cups should end up in the same position as they started. Participants were encouraged to explore strategies to optimize speed, performance, and learning during practice (Surkar et al., 2023).

#### 2.4.2 Intensive bout of Bimanual Training – Hand-arm Bimanual Intensive Therapy (HABIT)(Gardas et al., 2023; Surkar, Hoffman, Willett, et al., 2018)

Intensive bout of bimanual training involved six hours of HABIT per day for five consecutive days. HABIT is a well-established intervention shown to improve bimanual coordination in children with UCP. It was administered in a camp-based setting where participants engaged in structured, task-specific, bimanual activities in a playful context. Each participant was paired with 4 interventionists (i.e., occupational or physical therapy students), under the guidance and supervision of licensed PTs and OTs with at least 5-15 years of pediatric rehabilitation experience. The interventionists underwent training before the camp with an experienced researcher and clinician and treatment fidelity was ensured during the camp. Interventionists progressively increased the complexity of bimanual activities and graded the task demands based on the performance to allow the child to complete the tasks successfully. Throughout the HABIT camp, children were encouraged to use the affected and the less affected UE in a coordinated manner. Positive reinforcement and knowledge of performance were provided to motivate and reinforce desired goal-directed activities. Emphasis was placed on different roles of the affected UE, such as stabilizer, manipulator, and assistor depending on the child’s ability and task goal. Sessions were comprised of whole-task and part-task practice. Activities performed by the children were documented by the interventionists.

### 2.5 Data Acquisition

Participants underwent assessment of bimanual learning, spatiotemporal kinematics, and affected UE use during bimanual play at baseline, at visit 2 after short bout of bimanual training (75 trials of cup stacking), and at visit 8 after intensive bimanual training (HABIT).

#### 2.5.1 Bimanual learning

Participants performed three trials each of the three patterns of cup stacking shown in Figure 2. The time taken (in seconds) to complete each trial, or movement time, was recorded and nine trials were averaged for the performance measurement. Faster speed indicated better performance and enhanced bimanual learning.

#### 2.5.2 Kinematics of Bimanual Coordination

Two separate speed stack tasks were used to assess *bimanual coordination* (Figure 3A) and *symmetric performance* (Figure 3B) of the affected and less affected UEs. During testing, participants were seated in an adjustable chair in front of a table, with their back supported (erect), hips and knees at 90 degrees flexion, feet flat on the floor, elbows flexed at 90 degrees, and hands 30 cm apart at the edge of the table. For the bimanual coordination test, participants were instructed to use both hands cooperatively to remove cups from the stack and build a pyramid of 6 cups, following the pattern shown in Figure 3A. While performing the symmetric performance test, participants were asked to use both hands to move the cups concurrently at the target position (Figure 3B). During both tests, participants were encouraged to perform the tasks quickly and accurately using both hands on a ‘GO’ signal. One practice trial was provided before recording three trials for each test.

**Figure 3.**
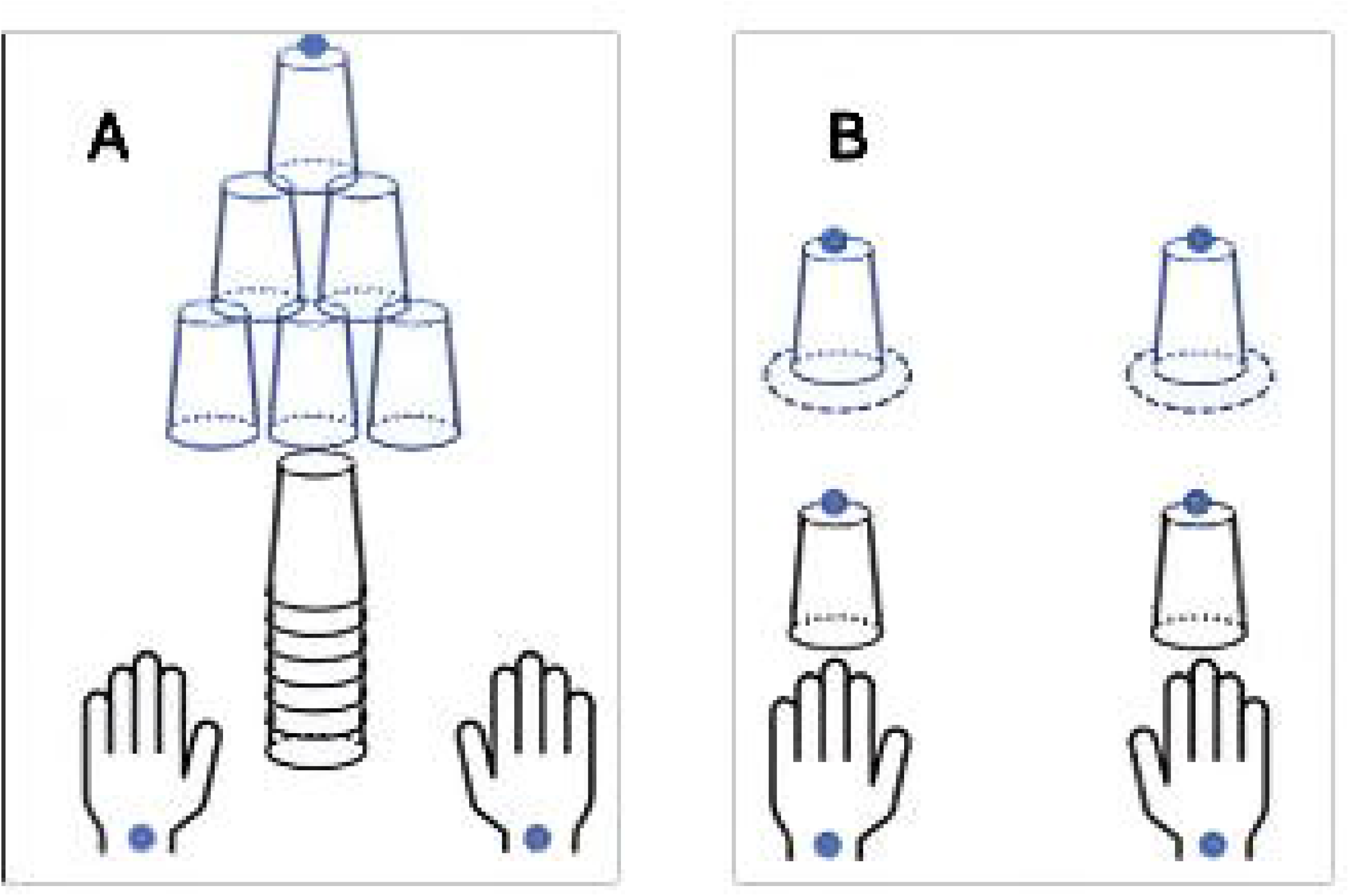
Speed stack task A. Bimanual coordination task: participants used both hands in coordination with each other to remove the cups from the stack and create a pyramid of six cups. B. Symmetric performance: participant used both hands to move the cups concurrently at the target position.

### 2.5.3 Spatiotemporal Kinematic Data Acquisition

3D kinematic data were collected during both bimanual coordination and symmetric performance tasks using a 15-camera Motion capture system (Qualisys AB, Gothenburg, Sweden). Reflective markers were placed on the midpoint of dorsal surface of the wrists and on the top of the final cup of the bimanual coordination task and on top of each cup for the symmetric task. Four markers were also placed on the corners of the table used for these tasks. Room calibration (space) was performed with set external x (medial-lateral axis), y (anterior-posterior axis), z (vertical axis) coordinates. All markers were digitized at a rate of 200 Hz and the digitized signals were processed through a dual low-pass digital (Butterworth) filter with a cutoff frequency of 6 Hz.

### 2.6 Outcome measures

#### 2.6.1 Bimanual learning

The average time (in seconds) required to complete each of nine trials of the cup stacking task. A reduction in movement time across trials is interpreted as an improvement in bimanual motor learning, reflecting increased efficiency and skill acquisition.

#### 2.6.2 Bimanual coordination task (Y. C. Hung et al., 2011, 2018; Simon-Martinez et al., 2020)

a. *Normalized movement overlap time:* The percentage of total task duration during which both arms are simultaneously active. An arm was considered active when the tangential velocity of its wrist marker exceeded 2.0 cm/s for a minimum of 100 milliseconds. This metric reflects the temporal overlap of bimanual engagement during task execution.
b. *Hand trajectory*: The total path length (in meters) traversed by each wrist marker during task execution. This measure reflects the spatial efficiency and movement economy of each hand throughout task completion.
c. *Total task duration:* The time (in seconds) from the movement onset of either hand to task completion, operationalized as the point at which cup marker speed drops below 2.0 cm/s and cup height exceeds 27 cm for at least 100 milliseconds. This measure captures the overall temporal span of task execution.
d. *Total participation time:* The cumulative duration (in seconds) following movement onset during which the tangential velocity of either wrist marker exceeds 2.0 cm/s. This measure reflects the total time either hand is actively engaged in the task.
e. *Peak tangential velocity:* The maximum tangential speed (in meters/second) of each wrist marker from movement onset until the corresponding cup becomes stationary. This measure represents the highest velocity attained by each hand during task execution.

#### 2.6.3 Symmetric performance task (Cacioppo et al., 2023)

a. *Task synchronization:* the temporal lag between the affected and less-affected arms at both movement onset and task completion. Movement onset was operationalized as the time point when the wrist marker’s tangential velocity exceeded 2.0 cm/s for a minimum of 100 milliseconds. This measure reflects the temporal alignment of bilateral hand actions during task execution.
b. *Task completion:* The duration (in seconds) from movement onset to the point at which each respective cup becomes stationary. This measure captures the total time required by each hand to complete the task independently.
c. *Peak tangential velocity:* The maximum speed (in meters/second) of each wrist marker from movement onset to the point at which the corresponding cup reaches a stable, stationary position.
d. *Hand trajectory: T*he total distance (in meters) traveled by each wrist marker from movement onset until the corresponding cup markers become stationary. This measure reflects the spatial path of hand movement during task execution.

#### 2.6.4 Assisting Hand Assessment (AHA) score

AHA is a reliable and valid outcome to assess the affected hand function and bimanual coordination, specifically in children with UCP. It consists of 20 items across six categories of arm use, with a rating scale from 1 to 4: 1 = no arm use, 4 = effective arm use. The raw AHA scores were converted into logit units (ranging from 0 to 100) for the final analysis. A change of 5 points on the AHA scale is regarded as clinically significant (Krumlinde-Sundholm, 2012; Krumlinde-Sundholm et al., 2007).

### 2.7 Statistical Plan

Data analyses were conducted using SPSS (version 28.0; IBM Corporation, Armonk, New York). We employed a general linear model (GLM) with repeated measures to analyze the effects of dose of training and extremity on the dependent variables (spatiotemporal kinematic variables; hand trajectory, total participation time and peak tangential velocity). Dose of training (baseline, short-bout, intensive bout of bimanual training) was considered a within-subject factor, and extremity (affected, non-affected) was considered a between-subject factor. We used a one-way repeated measures ANOVA to analyze the effects of training dose (within-subject factor) on the dependent variables: AHA scores, movement time, normalized movement overlap, total task duration, and task synchronization. Before conducting the analysis, we assessed the assumptions of the repeated-measures GLM. Mauchly’s test was used to evaluate the assumption of sphericity for the within-subject factor (dose), and the Greenhouse-Geisser correction was applied when sphericity was violated to adjust the degrees of freedom. The normality of residuals was checked using Q-Q plots, and the Shapiro-Wilk test was performed to detect any significant deviations from normality. Levene’s test was used to assess homogeneity of variances. The repeated-measures GLM was then used to examine the main effects of time and extremity, as well as their interaction. The significance level for the main effect of dose was set at α ≤ 0.05. Post-hoc Bonferroni pairwise comparisons were conducted to adjust for multiple comparisons between the dosages, with a significance level of α ≤ 0.01. In cases of a significant dose × extremity interaction, simple effects analyses were performed to further investigate differences across dose within each extremity. Partial eta-squared values were calculated to quantify the magnitude of significant findings.

## 3. Results

Twenty-eight children with UCP, age 6–16 years (mean ± SD: 10.4 ± 3 years, M = 22, F = 6) and Manual Ability Classification system levels I–III (I = 5, II = 9, and III = 14) participated in this study. Figure 4. illustrates the CONSORT diagram, detailing how participants progressed through the study, including withdrawals, and their inclusion in the analysis. Table 1 summarizes participant demographics and clinical characteristics.

**Figure 4.**
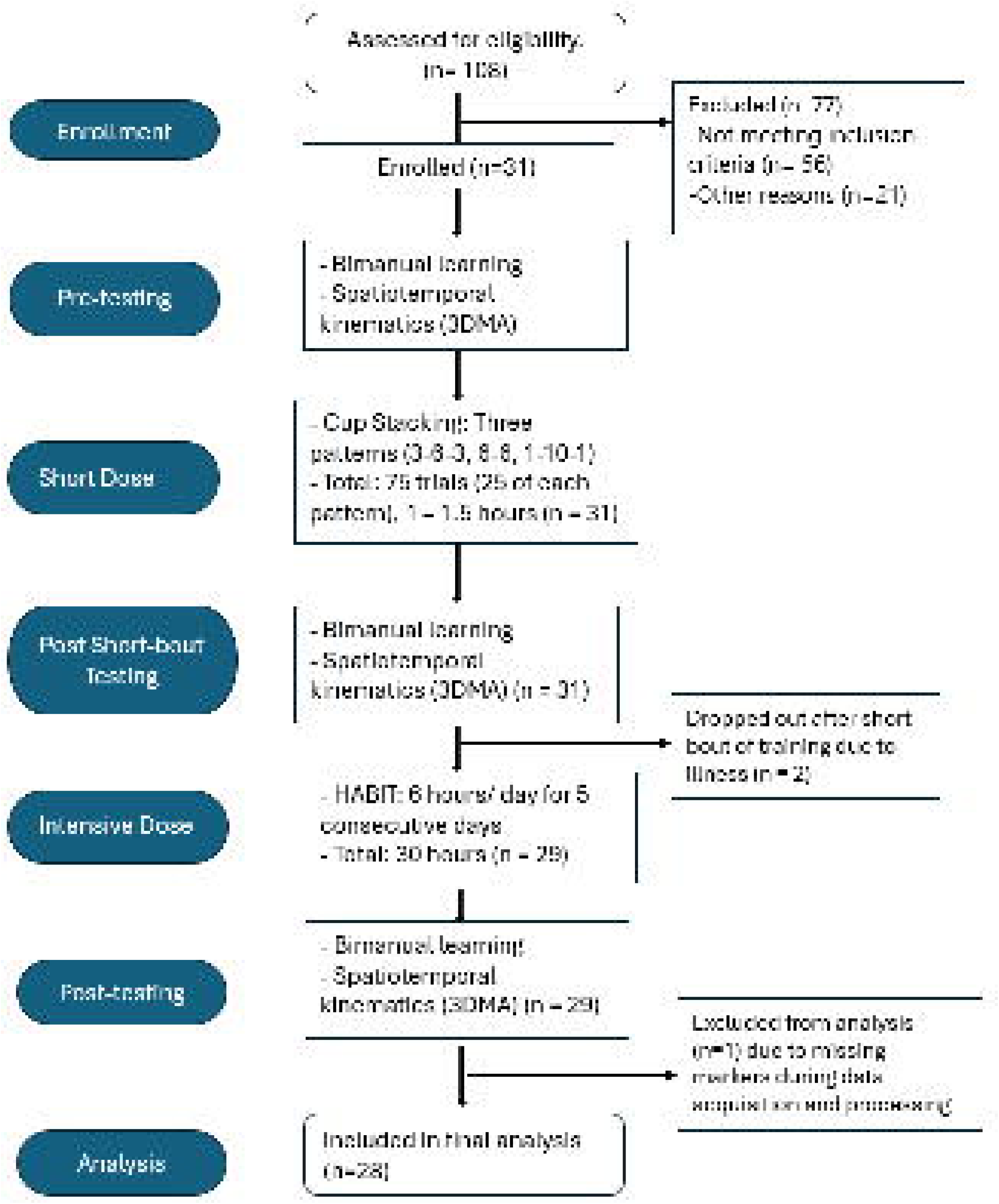
CONSORT diagram. A total of 108 participants were assessed for eligibility and 31 participants eventually participated and completed the short and intensive dose of bimanual training protocol. Of these, twenty-eight participants were included in the final analysis. Three participants were excluded: two withdrew after the short training dose due to illness, and one was excluded due to missing markers and tracking issues during kinematic data processing.

**Table 1.** Baseline demographic details of the study participants.

| Characteristics | Participants (n = 28) |
| --- | --- |
| Sex, n (%) |  |
| Male | 22 (78.6%) |
| Females | 6 (21.4%) |
| Age, mean (SD) | 10.4 (3) years |
| Side of hemiplegia, n (%) |  |
| Left | 12 (42.9%) |
| Right | 16 (57.1%) |
| Race, n (%) |  |
| White | 19 (67.9%) |
| African America | 4 (14.3%) |
| Asian | 3 (10.7%) |
| Multiracial | 2 (7.1%) |
| MACS levels, n (%) |  |
| I | 5 (17.9%) |
| II | 9 (32.1%) |
| III | 14 (50%) |

MACS indicates Manual ability classification.

### 3.1 Bimanual learning

There was a significant main effect of training dose on movement time (*F*_2, 44_ = 59.24, p < 0.001, partial η*^2^* = 0.73, Figure 5). Post-hoc pairwise comparisons with Bonferroni corrections showed significantly reduced movement time following both the short bout (mean difference = 9.85 seconds, p < 0.001, 95% confidence interval [CI] = 7.3 – 12.4), and intensive bout of training (mean difference = 11.7 seconds, p < 0.001, CI = 9.3 – 14.2) compared to baseline. No significant difference in movement time was observed between the short- and intensive training (mean difference = 1.9 seconds, p = 0.08, CI = -0.29 – 4.1), suggesting that both interventions were similarly effective in promoting bimanual motor learning. These results suggest that even brief training may yield meaningful improvements in clinical assessments of motor performance in children with UCP.

**Figure 5.**
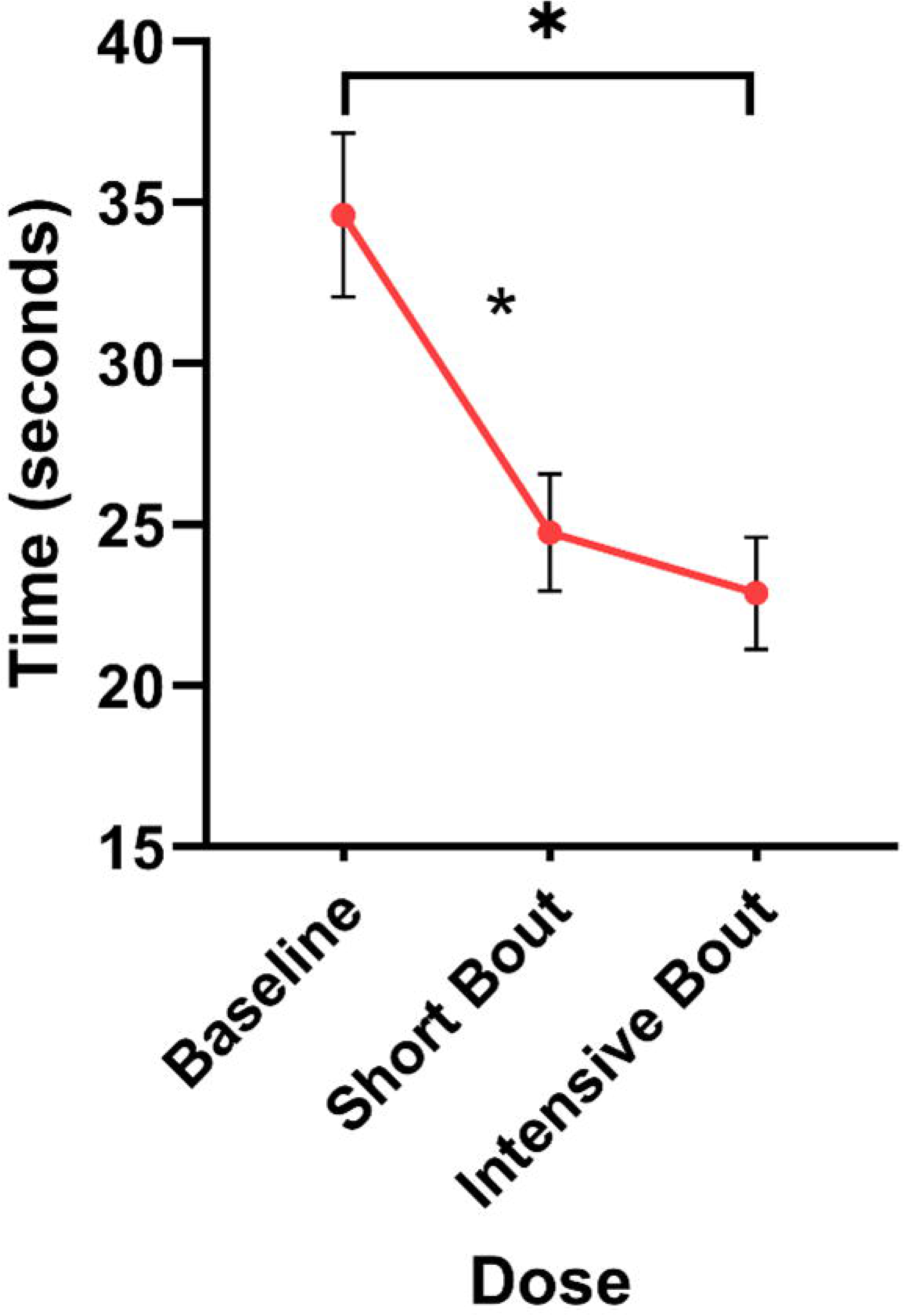
Effect of bimanual training dose on movement time in children with UCP: both short and intensive training significantly improved speed compared with baseline. Mean movement time scores (± SEM) are shown across three training dosages: baseline, short bout (75 cup stacking trials), and intensive bout (HABIT). Both training protocols resulted in significant reductions in movement time compared to baseline, indicating improved bimanual motor speed. The intensive bout showed a greater reduction than the short bout, although the difference did not reach statistical significance. *P ≤ .01, Bonferroni-corrected pairwise comparisons.

### 3.2 Spatiotemporal Kinematics – Bimanual Coordination Task

#### Normalized movement overlap

There was a significant main effect of training dose on normalized movement overlap (*F*_2, 54_ = 7.8, p = 0.001, partial η*²* = 0.22, Figure 6a). Post-hoc pairwise comparisons revealed a significant improvement in movement overlap following the intensive training bout compared to baseline (mean difference = 8%, p = 0.002, 95% CI = 3% – 13%). However, there was no significant increase in normalized movement overlap following the short bout of training compared to baseline (mean difference = 4.2%, p = 0.05, 95% CI = 0.1% – 8%), and between short and intensive bouts of training (mean difference = 3.7%, p = 0.02, 95% CI = 0.4% – 7%). These findings indicate the intensive training results in more robust improvements in bimanual engagement during task execution.

**Figure 6.**
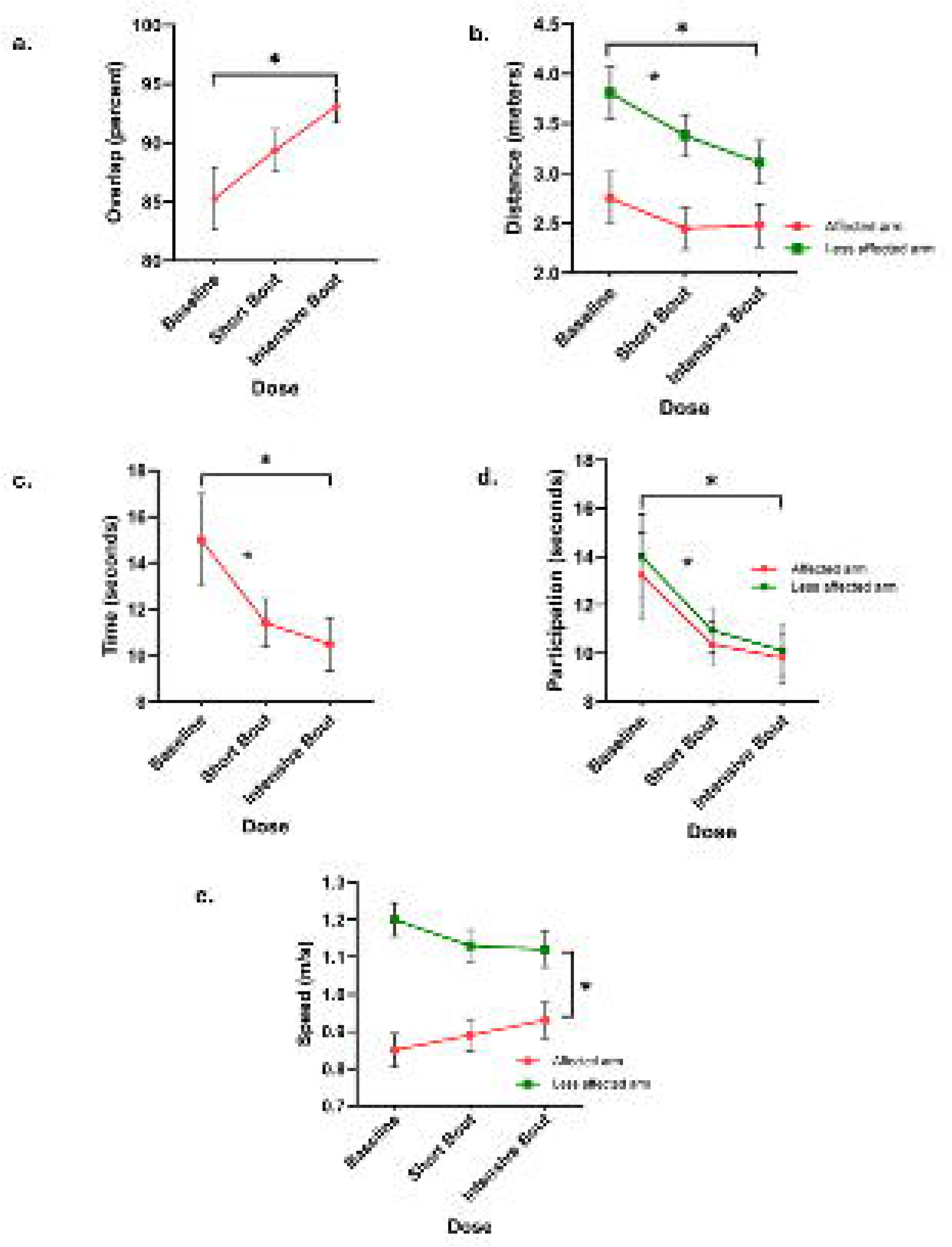
Effects of Bimanual Training Dose on Spatiotemporal Kinematics During a Bimanual Coordination Task. Mean ± SEM values for: (a) Normalized movement overlap, (b) Hand trajectory, (c)Total task duration, (d) Total participation time, and (e) Peak tangential velocity. Intensive training significantly increased movement overlap and reduced task duration, indicating enhanced coordination. Both training doses reduced total participation time and distance traveled. Peak tangential velocity of the affected arm increased significantly post-training, reflecting greater limb contribution. *p ≤ 0.01, Bonferroni-corrected.

#### Hand trajectory

The main effect of training dose on hand trajectory was significant (*F* (2, 108) = 3.56, p < 0.001, partial η² = 0.14; Figure 6b). Post-hoc pairwise comparisons showed that hand trajectory was significantly reduced following both the short bout and the intensive bout of training compared to baseline. Specifically, reductions were 0.37 m (p < 0.01, 95% CI: 0.09– 0.64) and 0.48 m (p < 0.001, 95% CI: 0.23 – 0.73), respectively. There was no significant difference between the short and intensive training bouts (mean difference = 0.12 m, p = 0.27, 95% CI: –0.09 – 0.33), indicating that both training protocols were equally effective in improving spatial efficiency and movement economy. No interaction was observed between training dose and extremity, suggesting that the effect of training on hand trajectory was consistent across both limbs.

#### Total task duration

There was a significant main effect of training dose on total task duration (*F*_2, 54_ = 12.75, p < 0.001, partial η*²* = 0.32, Figure 6c). Post-hoc pairwise comparisons with Bonferroni corrections showed that task duration was significantly reduced following both the short bout of training (mean difference = 3.6 seconds, p < 0.003, 95% CI = 1.3 – 5.9) and the intensive bout of training compared to baseline (mean difference = 4.52 seconds, p < 0.001, 95% CI = 2.3 – 6.7). These findings suggest that both training protocols enhanced task efficiency, likely reflecting improved speed and bimanual coordination. No significant difference in task duration was observed between the short- and intensive-bout of training (mean difference = 0.93 seconds, p = 0.09, 95% CI = -0.19 – 2), indicating that the two training protocols may be similarly effective in reducing the time required to complete the coordination task.

#### Total participation time

There was a significant main effect of training dose on total participation time (*F*_2, 108_ = 19.75, p < 0.001, partial η*²* = 0.27, Figure 6d). Post-hoc pairwise comparisons showed significant reductions in participation time following both the short training bout (mean difference = 2.9 seconds, p < 0.001, 95% CI = 1.5 – 4.4]) and the intensive training (mean difference = 3.7 seconds, p < 0.001, 95% CI = 2.3 – 5.04]) relative to baseline. These findings indicate a reduction in total participation time following both training doses compared to baseline. A greater reduction in participation time after the intensive bout suggests that a higher dose results in more efficient movement patterns compared to the short bout. These reductions suggest improved movement efficiency during the bimanual coordination task. No significant difference in participation time was observed between the short- and intensive-bout of training (mean difference = 0.67 seconds, p = 0.07, 95% CI = -0.04 – 1.4), indicating that both training protocols may yield comparable benefits. No interaction effect was found between training conditions and extremities, suggesting that the effects of training on participation time were consistent across both upper limbs.

#### Peak tangential velocity

A significant interaction was observed between training dose and limb (F (2,108) = 3.64, *p* < 0.03, partial η² = 0.06; Figure 6e), indicating that the effects of training on peak tangential velocity differed between the affected and non-affected UEs. The affected UE’s peak tangential velocity increased more than the non-affected following both the training doses. There was also a robust main effect of extremity (mean difference = 0.26 m/s, *p* < 0.001, 95% CI [0.15, 0.38]), confirming that peak tangential velocities were substantially different between the limbs. In contrast, the main effect of training dose on peak tangential velocity was not significant (F (2,108) = 0.12, *p* = 0.88, partial η² = 0.002). A follow-up post hoc paired *t*-tests revealed that the affected limb demonstrated greater and more consistent improvements in peak tangential velocity across all training comparisons: 0.04 m/s (baseline to short dose), 0.04 m/s (short to intensive dose), and 0.08 m/s (baseline to post-intensive). In contrast, the non-affected limb exhibited smaller and more variable changes: −0.06 m/s, −0.08 m/s, and −0.01 m/s for the same respective comparisons. These findings suggest that the affected extremity exhibited more favorable and systematic gains in movement speed, particularly in response to intensive bimanual training.

### 3.3 Spatiotemporal Kinematics – Symmetric Performance Task

#### Task synchronization

There was no significant effect of training dose on task synchronization (*F*_2,104_ = 1.01, p = 0.37, Figure 7a), indicating that the time lag between the extremities during onset and task completion remained similar across different doses of bimanual training for symmetric performance task.

**Figure 7.**
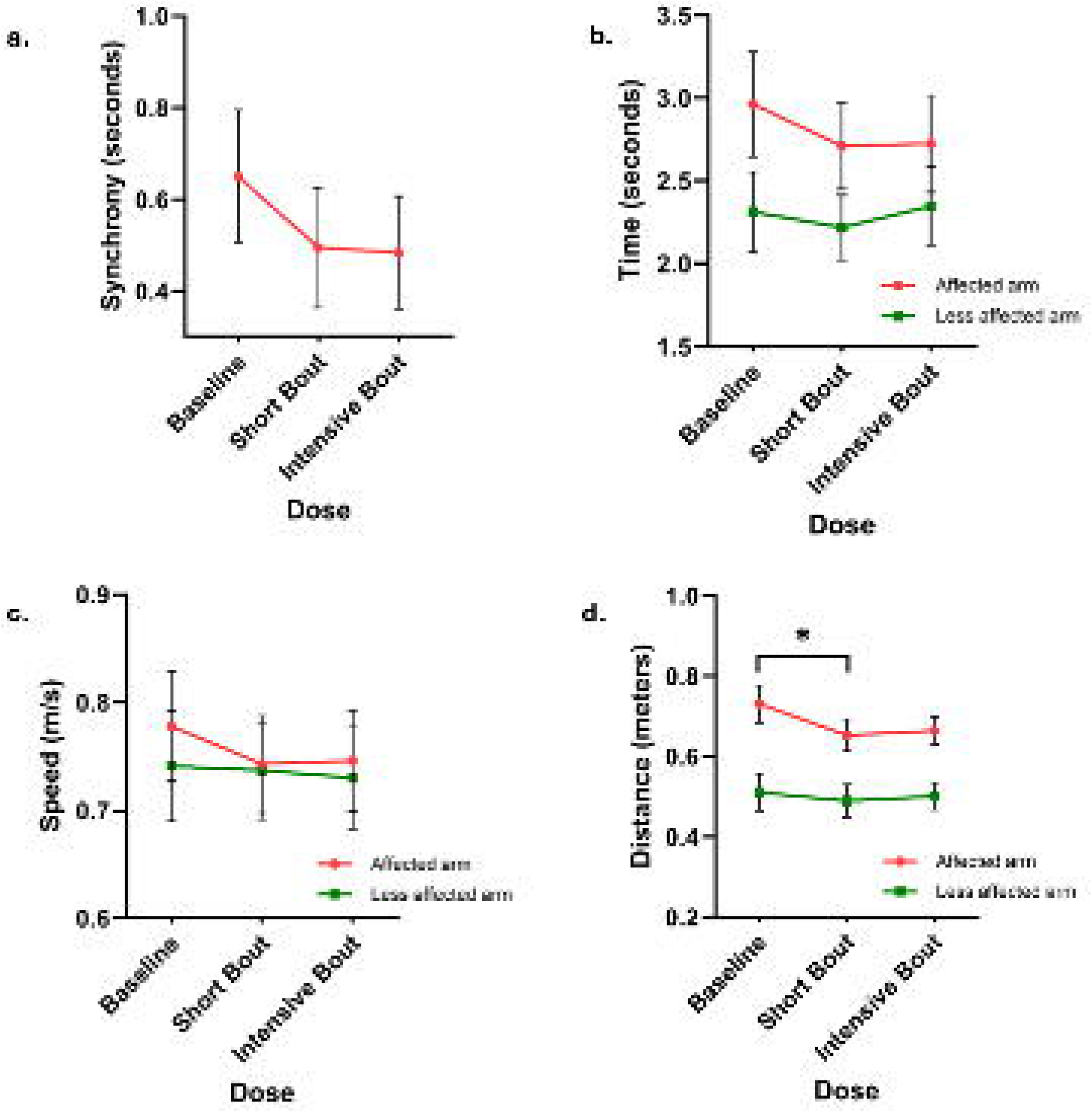
Effects of Bimanual Training Dose on Spatiotemporal Kinematics During a Symmetric Performance Task. Mean ± SEM values for: a) Task synchronization, b) Task completion time, c) Peak tangential velocity, and d) Hand trajectory. Hand trajectory significantly reduced after a short bout of training, suggesting improved efficiency of the affected arm’s movement. No significant changes were observed in all other variables across both the training doses.

#### Task completion time

The main effect of dose of training was not significant (*F*_2,104_ = 1.46, p = 0.24, Figure 7b). No significant interaction effect (*F*_2,104_ = 0.90, p = 0.41) was observed between training dose and extremities, suggesting that varying doses of bimanual training did not differentially influence task completion time for either extremity during the performance of a symmetric bimanual task.

#### Peak tangential velocity

The main effect of training dose on peak tangential velocity was not significant (*F*_2,104_ = 0.39, p = 0.68, Figure 7c). No interaction effect (*F*_2,104_ = 0.17, p = 0.84) was observed between training dose and extremities, indicating that varying doses of bimanual training did not differentially affect peak tangential velocity in either the affected or unaffected upper extremities.

#### Hand trajectory

There was a significant main effect of training dose (*F*_2,104_ = 3.59, p < 0.03, partial *^2^*= 0.07, Figure 7d). Post-hoc pairwise comparisons revealed that the hand trajectory was significantly reduced after short bout of training compared to baseline (mean difference = 0.05 meters, p = 0.005, 95% CI = 0.02 – 0.08). No significant difference in hand trajectory was observed between short and intensive bout of training (mean difference = 0.01 meters, p = 0.55, 95% CI = 0.03 – 0.05). No significant interaction effect was found between training dose and extremities.

### 3.4 Changes in Assisting Hand Assessment Scores

There was a significant main effect of training dose on AHA logit units [p < 0.001, Figure 8a]. Post-hoc pairwise comparisons using Bonferroni corrections showed statistically significant increase in the AHA scores following both the short bout (mean difference = 1.36, p < 0.001, 95% confidence interval [CI] = 0.74 – 1.9), and intensive bout of training (mean difference = 7.27, p < 0.001, CI = 5.14 – 9.4) compared to baseline. Additionally, AHA scores were significantly higher following intensive compared to short bout of training (mean difference = 5.9, p < 0.001, CI = 4.1 – 7.7), indicating a dose-response relationship favoring the more intensive bimanual training. Importantly, only the intensive training exceeded the minimal clinically important difference (MCID) for the AHA (5 logit units), yielding a clinically meaningful gains (mean change = 7.3 logits), whereas the short bout training (mean change = 1.3 logits) did not. These findings suggest that higher training intensity may lead to greater functional use of the affected hand during bimanual coordination activities.

**Figure 8.**
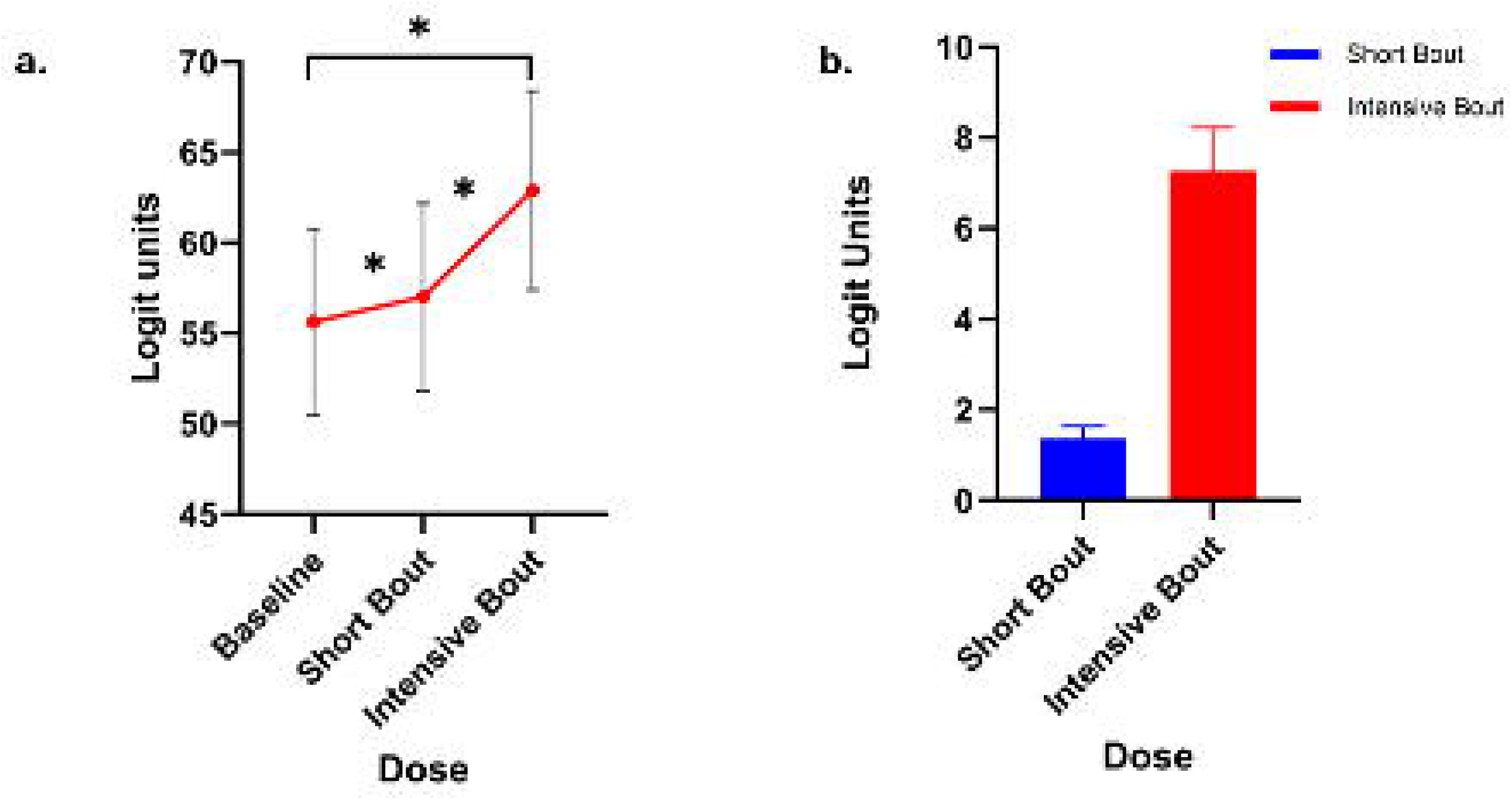
**a**. Effects of Bimanual Training Doses on AHA logit units. Values represent mean scores of AHA logit units (mean ± SE). The results showed significant improvements in affected hand function and bimanual coordination after short and intensive bout of training. **b**. Changes in the AHA logit scores compared to baseline. Values represent AHA logit change scores from baseline (mean ± SE). The results show that the intensive training dose led to significantly greater and clinically meaningful improvements in the use of the affected upper extremity during bimanual activities compared to the short training dose.

## 4. Discussion

The primary purpose of this study was to compare two doses of bimanual training (short and intensive bouts) on bimanual learning and spatiotemporal characteristics of bimanual coordination in children with UCP. We employed a novel speed stack task under two paradigms– bimanual coordination and symmetric performance– to examine changes in spatiotemporal characteristics following training. Children with UCP showed improved bimanual learning after both training doses, evidenced by reduced movement times. In bimanual coordination task, the intensive bout notably increased movement overlap and reduced task duration suggesting enhanced bimanual coordination. Furthermore, both doses decreased participation time and hand trajectory length, indicating greater efficiency, and increased peak velocity of the more affected limb, reflecting improved more affected limb use. Contrary to our hypothesis, minimal changes were observed during the symmetric task. Overall, these findings underscore the importance of training dose and task specificity in improving spatiotemporal aspects of bimanual learning in children with UCP.

### 4.1 Motor Learning Occurs Even with Short bout of Bimanual Training

Impairments in bimanual coordination resulting from unimanual deficits in children with UCP present a major challenge for therapists, often leading to functional dependence and reduced quality of life (Cacioppo et al., 2023). The ability to perform bimanual tasks more efficiently following training serves as an indicator of improved motor learning (Y. C. Hung & Gordon, 2013). In our study, movement time (i.e., the average time taken to complete nine trials of the speed stacking task) decreased by 28.6% after the short bout and by 34.1% after the intensive bout, suggesting that both training doses enhanced movement speed reflecting bimanual learning. The intensive bout led to greater improvements, likely due to the higher training dose, structured practice, and increased problem-solving demands during the 30 hours of HABIT. These findings align with previous studies showing that intensive UE protocols, such as CIMT and HABIT, result in greater improvements in UE function and bilateral hand use (Sakzewski et al., 2014, 2015). A closely related study also demonstrated that 15 days of speed stacking (three trials per day) enhanced motor learning and movement speed in children with CP (Y. C. Hung & Gordon, 2013). Similarly, a 5-week (15-hour) task-oriented intervention improved motor learning during a reach and grasp task (Robert et al., 2013). Interestingly, even the brief training bout (1–1.5 hours) of training involving 75 trials of speed stacking across three patterns, produced substantial gains in bimanual learning. These improvements may be attributed to bimanual adaptations driven by the task-specific, high-repetitive nature of the speed stacking practice, consistent with established motor learning principles (Muratori et al., 2013). Importantly, the duration of the short-bout training closely aligns with that of a typical outpatient therapy session for children with UCP. The meaningful gains observed following this brief training dose suggest that the motor system in children with UCP retains the capacity for rapid, within-session adaptation and within standard clinical timeframes, task-specific, high-repetition bimanual practice can lead to significant functional improvements in this population.

### 4.2 Dose-Dependent Enhancement of Interlimb Coordination and Hand Trajectory Through Task-Specific Bimanual Training

Children with UCP demonstrated significant dose-responsive improvements in interlimb coordination, as evidenced by increased movement overlap during a complex, ecologically valid bimanual task. Movement overlap, quantifying the proportion of time both hands move concurrently, increased by 4.7% following a short training bout and by 9.5% following intensive training. These findings suggest that both low and high doses of structured, task-specific practice can effectively modulate temporal coupling between limbs, a core deficit in UCP that impairs functional bimanual performance (Y. C. Hung et al., 2011).

The observed gains reflect underlying improvements in the temporal synchrony of bimanual actions, which are critically dependent on the integrity of interhemispheric communication (MacDonald et al., 2021). Children with UCP often exhibit disrupted development of transcallosal and corticospinal pathways, leading to impaired interlimb timing (Simon-Martinez et al., 2022). The greater gains in movement overlap following the intensive 30-hour intervention point to the importance of repetition and task complexity in engaging these distributed motor control networks and driving experience-dependent plasticity.

Compared to prior reports of larger improvements—such as an 18% increase after 90 hours of HABIT (Y. C. Hung et al., 2011) – the more modest gains observed here may reflect the older participant cohort, shorter training duration, and the use of a more challenging, sequential bimanual task. Nevertheless, the fact that even a single short training session yielded measurable improvements underscores the brain’s responsiveness to targeted motor learning and highlights the potential of low-dose interventions to initiate meaningful changes in coordination.

In parallel, hand trajectory, defined as the total distance traversed by each hand during task performance, revealed dose-dependent refinements in movement efficiency. Both training bouts led to reductions in hand path length, with the intensive bout eliciting a larger decrease (14.9%) compared to the short bout (11.3%) training. This reduction likely reflects streamlined motor execution and reduced extraneous motion, aligning with more efficient neural representations of the task (Simon-Martinez et al., 2020; Surkar et al., 2019). Intriguingly, the affected UE exhibited a ∼3% increase in trajectory distance, while the less affected UE showed an ∼8% decrease following the intensive training. This asymmetric pattern suggests a redistribution of task demands, wherein the affected limb contributes more actively to task execution, thereby fostering balanced interlimb coordination.

These kinematic patterns align with earlier findings by Simon Martinez et al., who reported reduced hand trajectory in the affected UE following CIMT during simpler reaching and grasping tasks. Simon-Martinez et al., 2020). Despite methodological differences, our use of a complex, ecologically valid speed-stacking task allowed for the detection of similar neural and behavioral adaptations, even after shorter training durations. Collectively, these results underscore the dual importance of training dose and task specificity in driving meaningful improvements in motor behavior. They also highlight the clinical utility of detailed kinematic assessments in capturing nuanced changes in coordination, particularly those that reflect enhanced integration and efficiency of the affected UE during functionally relevant bimanual activities.

### 4.3 Training-Induced Modulation of Speed in Bimanual Coordination

Both short and intensive bouts of bimanual training yielded measurable improvements in motor execution efficiency, as reflected by reduced total task duration and total participation time, and increased velocity during a complex bimanual coordination task.

Training yielded a marked reduction in total task duration—a proxy for movement speed—with a 24.0% decrease following the short training bout and a 30.1% decrease following the intensive bout, relative to baseline performance. The greater improvement observed after intensive training likely reflects enhanced interlimb coordination and more robust consolidation of bimanual motor learning, attributable to the higher training dosage (Y. C. Hung et al., 2011, 2018). Importantly, the short-duration intervention, comprising 75 repetitions of the speed-stacking task, also resulted in a clinically meaningful reduction in task completion time, underscoring the potency of even brief, task-specific practice. These results reinforce the therapeutic value of structured, outcome-driven motor training in pediatric neurorehabilitation and advocate for the strategic integration of task intensity in intervention design.

Total participation time, defined as the cumulative duration post-movement onset during which either hand exceeds a speed threshold of 2 cm/s, decreased by 22.1% following the short bout and by 27.2% after intensive training. Notably, this reduction was symmetric across both UEs, suggesting improved temporal alignment and cooperative engagement of both limbs. These findings parallel the observed reductions in overall movement time, reinforcing the interpretation that even brief bouts of task-specific practice can refine temporal coupling and accelerate task execution in children with UCP.

Our findings revealed divergent patterns in peak tangential velocity between the two UEs across both training intensities. Peak tangential velocity—defined as the peak speed of each hand from movement initiation to task completion—decreased progressively in the less affected UE but increased in the more affected UE. This asymmetry likely reflects a compensatory shift in motor contributions, with improved neuromuscular engagement of the more affected limb during the bimanual coordination task. These observations are consistent with prior work by Gordon et al. (Y. C. Hung et al., 2018; Y. C. Hung & Gordon, 2013), and Simon Martinez et al., (Simon-Martinez et al., 2020) who similarly reported enhanced motor performance of the more impaired limb following intensive task-specific training. We hypothesize that the increased speed of the more affected UE facilitated the overall reduction in task duration, increased hand path trajectory, and improved interlimb synchrony. Collectively, these changes in movement dynamics underscore the clinical utility of both short and intensive training protocols in optimizing temporal efficiency and promoting functional integration of the more affected limb during complex bimanual activities.

### 4.4 Symmetric Task Performance and Limited Differential Gains in Bimanual Coordination

In the present study, we employed a symmetric bimanual performance task in which participants simultaneously grasped and placed two cups into spatially aligned target zones. This task was designed to elicit in-phase, temporally coupled movements of both upper extremities (UEs), paralleling bilateral simultaneous paradigms used in prior kinematic assessments of bimanual coordination in children with UCP (Langan et al., 2010; Utley & Sugden, 1998). Our results demonstrated comparable reductions in temporal asynchrony following both the short (24.6%) and intensive (24.2%) training bouts, relative to baseline. These reductions suggest enhanced bilateral coupling and synchronization, likely mediated by improved interhemispheric communication and reduced sensorimotor delay (Forssberg, 1999). However, the absence of a dose-dependent effect highlights the possibility of an early saturation of gains in symmetric coordination, particularly in tasks relying on highly coupled in-phase movement patterns.

Task completion time showed only marginal improvements (∼4%) across both training doses, and no significant inter-dose differences. The more affected UE consistently required longer to complete the task (∼2.8 seconds vs. ∼2.3 seconds for the less affected UE), yet it paradoxically exhibited slightly higher peak tangential velocity (0.76 m/s vs. 0.74 m/s). This may reflect reduced motor control and corrective adjustments, as higher speed in the presence of delayed completion time suggests inefficient or less accurate movement execution (Langan et al., 2010; Utley & Sugden, 1998). Similarly, the hand trajectory of the more affected UE remained longer than that of the less affected UE, despite post-training reductions of 8.1% and 6.5% following the short and intensive bouts, respectively. The dissociation between speed and accuracy metrics, especially in the more affected UE, is likely underpinned by impaired sensorimotor integration and mirror movements (Rudisch et al., 2016). These involuntary, mirrored activations of the contralateral limb are frequently observed during symmetric bimanual tasks and tend to emerge more prominently in the less affected UE (Rudisch et al., 2016). Their presence can obscure the distinct contributions of each limb and reduce task specificity, leading to homogenized performance patterns that limit the detection of dose-responsive improvements. Furthermore, prior studies suggest that during symmetric tasks, the more affected limb often initiates movement, with the less affected limb adjusting its timing and kinematics to achieve simultaneous task execution (Langan et al., 2010). This compensatory strategy may mask unilateral impairments and create an artificial symmetry that reduces task sensitivity. Lastly, the brief duration of the symmetric task (∼2–3 seconds) likely limited the resolution with which nuanced changes in coordination could be captured. High-speed execution over a short time window may not allow sufficient opportunity for adaptive motor strategies to manifest, particularly in metrics such as trajectory optimization or interlimb decoupling. As such, the symmetric task used in this study may have lacked the sensitivity required to detect more subtle training-induced refinements in motor control, especially those emerging after intensive practice.

### 4.5 Task-Specific Generalization and Transfer

The use of two distinct tasks—a coordinated bimanual task and a symmetric performance task— allowed us to explore how improvements generalize across task contexts. While both tasks required two-handed control, they differed in complexity, pacing, and degrees of bimanual asymmetry. Notably, improvements in movement synchrony and trajectory were more prominent in the bimanual coordination task, especially following intensive training. This suggests that more complex, asymmetric bimanual tasks may be more sensitive to changes in coordination and motor planning and may benefit more from high-dose training protocols. Conversely, the symmetric task, which involved relatively simpler and more mirrored actions, appeared to benefit modestly from short training but showed limited sensitivity to dose-dependent effects. This differential responsiveness may be explained through task-specific motor learning principles—whereby improvements are most pronounced in tasks closely aligned with the practiced activity. The degree to which motor skills transfer across tasks is thought to depend on the similarity of task demands, neural circuitry, and sensory feedback mechanisms (Leech et al., 2022; Schaefer et al., 2013; Sullivan et al., 2008). The stronger response in the bimanual coordination task underscores the need to include complex, functionally relevant tasks in training to maximize generalization and cortical engagement.

### 4.6 Dose-dependent gains in functional use and kinematic precision

Intensive bimanual training led to clinically meaningful improvements in upper limb function in children with UCP, with AHA scores improving by >5 logits. In contrast, short-bout training produced minimal gains (1.3 logits), confirming a dose-dependent effect. Three-dimensional motion analysis revealed that even brief training (∼75 repetitions) elicited measurable improvements in spatiotemporal control—enhanced hand cooperation (increased movement overlap), faster performance (shorter task duration), and greater motor efficiency (higher peak velocity, reduced path length). These kinematic changes, undetectable by observational scales, offer mechanistic insights into early motor adaptations. Our findings underscore the synergistic role of training dose and task specificity and highlight the utility of combining clinical and instrumented measures in pediatric neurorehabilitation research.

### 4.7 Study Limitations and Directions for Future Research

Few limitations of this study warrant consideration. First, operational definitions of select kinematic variables, such as peak tangential velocity, may benefit from further refinement. While peak velocity is frequently employed as an index of motor performance, it reflects only the instantaneous maximum speed of the limb and may not fully capture the efficiency or consistency of movement. Calculating average velocity across the full duration of the task may yield a more holistic representation of motor output and its contribution to overall task performance. Second, this study did not stratify outcomes based on the severity of hand impairment or other participant characteristics such as age, sex, or lesion type, which may have influenced individual responses to training. Future studies should incorporate these variables to facilitate a more nuanced understanding of treatment responsiveness and to guide the development of personalized, precision-based rehabilitation interventions. Lastly, prior investigation incorporated joint angle kinematics following training and reported alterations in kinematics post-CIMT during simpler unimanual reaching and grasping tasks. Future studies should evaluate joint kinematics during ecologically valid, multistep activities such as speed stacking, which could offer deeper insights into how training-induced changes in motor control translate to functionally relevant, bimanual behaviors. Incorporating such analyses may help refine upper extremity rehabilitation strategies to better support real-world task execution.

### 4.8 Conclusion

In children with UCP, both short and intensive bouts of bimanual training improved bimanual learning and spatiotemporal aspects of bimanual coordination, with the intensive dose yielding significantly greater gains. Notably, our findings demonstrate that even a brief period of training using an ecologically valid task, such as speed stacking, can enhance spatiotemporal coordination. However, only the intensive training dose resulted in clinically meaningful improvements in AHA logit scores, highlighting the sensitivity of objective kinematic measures, such as 3DMA, in detecting subtle changes in bimanual coordination that may not be captured through observational assessments alone. Collectively, these results emphasize the importance of optimizing training dosage, employing task-specific interventions, and incorporating precise assessment tools to accurately evaluate the efficacy of bimanual training in children with UCP.

## Acknowledgments

We thank the children and their families who participated in the study, certified clinical therapists (Drs. Christine Lysaght and Caroline Adams), and all the volunteer (physical and occupational therapy students) interventionists for their contributions to the HABIT camp. We sincerely thank the graduate research assistants Brody Morton, Taylor Key, Caroline Pusey-Brown, Katie Woosley, Natalie McBryde, Grant Kirkman, and Tanner Hallgren for their contributions to this project.

## Declaration of competing interest

The authors report there are no competing interests to declare.

## CRediT authorship contribution statement

**Shailesh S. Gardas:** Writing – review & editing, Writing – original draft, Software, Visualization, Software, Methodology, Investigation, Formal Analysis, Data Curation. **John Willson:** Writing – review & editing, Validation, Supervision, Software, Methodology, Investigation. **Swati M. Surkar:** Writing – original draft, Writing **–** review & editing, Conceptualization, Visualization, Validation, Software, Resources, Project administration, Methodology, Investigation, Funding acquisition, Formal Analysis, Data curation.

## Funding

This work was supported by the Eunice Kennedy Shriver National Institute of Child Health & Human Development of the National Institutes of Health under Award Number R03HD107644 to PI, Swati M. Surkar, and APTA’s Pediatric Physical Therapy (Grant No. 21-0810) to PI, Swati M. Surkar.

## Data availability

Data will be made available on request.

## Notes

### Competing Interest Statement

The authors have declared no competing interest.

### Clinical Trial

NCT05355883

### Author Declarations

Ethics committee/IRB of East Carolina University gave ethical approval for this work

